# Benefit incidence analysis of decentralized Truenat MTB Plus and MTB-RIF Dx compared to hub-and-spoke Xpert MTB/RIF in Mozambique and Tanzania (TB-CAPT CORE trial)

**DOI:** 10.1101/2025.10.20.25338356

**Authors:** Lelisa Fekadu Assebe, Saima Bashir, Akash Malhotra, Délio Elísio, Antonio Machiana, Anange Lwilla, Jerry Hella, Neenah Young, Mikaela Watson, Vinzeigh N. Leukes, Adam Penn-Nicholson, Morten Ruhwald, Berra Erkosar, Leyla Larsson, Katharina Kranzer, David Dowdy, Claudia M. Denkinger, Manuela De Allegri, the TB-CAPT Consortium

## Abstract

**Introduction:** Tuberculosis (TB) is a major public health threat worldwide and about one-third of cases go undiagnosed or unreported, leading to continued transmission of the disease. While point-of-care diagnostics are crucial to improve case detection, their distributional financial impact remains underexplored. Understanding how the costs and benefits of these tools are distributed across socio-economic groups is critical to ensure equitable access. This study aims to estimate the benefit incidence of implementing Truenat MTB Plus and Truenat MTB-RIF Dx at point-of-care (intervention arm), compared to the hub-and-spoke Cepheid Xpert MTB/RIF (‘Xpert‘) system or onsite sputum-smear microscopy test (control arm), considering utilisation across the wealth quintiles in Mozambique and Tanzania.

**Methods:** We conduct a benefit incidence analysis (BIA) using data derived from the TB-CAPT CORE multi-center, cluster randomized controlled trial with a sub-study on patient and provider costs. Our main outcome measure was diagnosis and TB treatment initiation within 7 days of diagnosis. Both patient and health system costs were included in our analysis. We estimated the benefit incidence from health system or societal perspective (combining both patient and health system) for each TB diagnostic strategy reported across wealth quintiles. We constructed concentration curves to visualize the cumulative share of benefit incidence distribution and estimate indices to quantity the magnitude of inequality in benefit for the trial arm and country.

**Result:** The proportion of people diagnosed with TB and started on treatment was significantly higher among the poorest quintiles (28 patients (25%) in Mozambique and 35 patients (27%) in Tanzania) compared to the least poor quintiles (20 patients (18%) in Mozambique and 15 patients (12%) in Tanzania). A total of 147 individuals in the intervention group initiated TB treatment within 7 days, compared to 95 in the control group. This reflects the intervention arm had more than 1.5 times higher rates of TB treatment initiation compared to control arm. This difference was especially notable in Mozambique. While the societal cost of TB treatment initiation was higher in the intervention arm as compared to the control arm, the public benefit for treatment initiation was more beneficial to poorer populations, with the benefit incidence slightly skewed towards the poor (concentration index of –0.0816 (confidence interval: – 0.0829: – 0.0804)).

**Conclusion:** The decentralized Point-of-care (POC) testing substantially improved TB treatment initiation within 7 days compared to standard care, ensuring that the investment reached those who need it most, particularly the poorest population subgroups. These finding showed a pro-poor distribution of benefit incidence emphasizing the importance of decentralized POC to improve diagnostic access and promote equity.

## Background

Tuberculosis (TB) remains a critical global health issue, with substantial morbidity and mortality rates. In 2023, TB was responsible for approximately 10.8 million cases and 1.25 million deaths worldwide [1]. A major factor contributing to this high morbidity and mortality is the gap in TB detection, with about one-third of the estimated TB cases not being diagnosed and reported [2]. This issue is driven in parts by inadequate access to more sensitive TB diagnostic tests in decentralized settings where most patients present [3].

In response to these challenges, many high TB burden countries continue to rely on smear microscopy in peripheral health facilities or have established specimen referral networks linking peripheral health facilities (spokes) to district-level hubs equipped with GeneXpert (Cepheid, Sunnyvale, CA, USA) machines for testing with Xpert or Xpert Ultra MTB/RIF (‘Xpert’). Such system enables the effective use of costly diagnostic technologies that are strategically placed at appropriate tiers of the laboratory system. Even though the hub-and-spoke model enhances access to Xpert testing, it faces several challenges. This includes difficult transportation of biological specimens, compromised standard biosafety and bio-security requirements, long turn-around-times for transportation and result delivery, limited accessibility of the courier system to remote and, hard to reach health facilities, and inadequate management capacities at national, regional, referring and referral testing facility [4-7].

The introduction of decentralized diagnostic tools, such as the Truenat MTB Plus and Truenat MTB-RIF diagnosis at peripheral facilities or clinics, offers potential solutions by placing diagnostic capabilities closer to patients [8, 9]. Such diagnostic platforms produce results in one hour at similar sensitivity to Xpert [10, 11]. Following diagnosis, prompt and effective treatment is essential to maximize the impact on reducing mortality and the spread of TB [12]. Concurrently, the WHO has endorsed centralized diagnostic platforms that enable multi-disease testing (e.g., TB, HIV, hepatitis) and high-throughput capabilities, potentially providing cost savings, improving system efficiency, and expanding patient access [13, 14]. An assessment of strategies for developing specimen referral networks using both centralized and decentralized systems revealed mixed results, demonstrating effectiveness of both strategies dependent on context [15].

These controversies and technology advancement require a critical evaluation of whether to adopt a centralized or decentralized diagnostic strategy for TB [16]. To make an informed decision, it is necessary to analyze not only the clinical effectiveness and cost effectiveness but also evaluate the distribution of financial resources invested in the point-of-care diagnostic tool across socio-economic groups, particularly benefiting underserved populations and reducing health inequalities. Mozambique and Tanzania have high TB burden and face barriers to timely diagnosis and treatment, particularly among underserved populations. Health system constraints and centralized diagnostic services contribute to inequities in access, undermining TB control efforts. A Benefit Incidence Analysis of whether investments in decentralized point-of-care testing reach the most disadvantaged population is crucial for supporting equitable resource allocation. Although studies in this area are limited, evidence from Enugu State, Nigeria, shows that TB treatment benefits tend to be pro-poor but remain concentrated in urban areas, underscoring persistent inequities and the need to improve access for rural populations [17]. This study builds on the TB-CAPT Core trial, a pragmatic, cluster-randomized controlled trial employed in Tanzania and Mozambique. The objective of this study was to assess the benefit incidence of delivering TB diagnostics through various delivery approaches in both countries.

## Methods

### Study setting and design

The TB CAPT core trial employed a repeated cross-sectional study design to collect primary data from 29 peripheral health facilities located across four participating institutions in Tanzania and Mozambique. The data collection period was from August 2022 – June 2023. In this study, we estimated the benefit of TB diagnosis and early treatment initiation by comparing two diagnostic delivery approaches. The intervention arm involved onsite Truenat testing using Truenat MTB Plus and Truenat MTB-RIF Dx at point-of-care facilities. The control arm used a hub-and-spoke model with samples referred to centralized laboratories for Cepheid X-pert MTB/RIF (X-pert) testing or parallel onsite sputum-smear microscopy. The health facilities were randomly allocated to the intervention arm, comprising 15 health facilities, and control arm, consisting of 14 health facilities [16].

### Sampling and data collection procedures

We used data collected from the TB-CAPT core trial survey [18], as well as from subsample study on patient and health system costs, as the basis for our benefit-incidence analysis (BIA). The TB-CAPT survey included a representative sample of 3,987 participants, capturing comprehensive information on participants’ background characteristics, epidemiological and diagnostic outcomes, health status, and asset ownership.

To estimate patient costs, we employed a systematic random sampling of every tenth TB CAPT survey participant (i.e., 3,987) to select 388 samples (approximately 10%) across the 29 participating clinics. We applied blocked randomization within four strata to ensure balanced representation across countries and regions. Additionally, we collected health system costs data from 19 out of 29 health facilities of the TB-CAPT core trial study sites through applying a bottom-up costing approach [19]. The subsample survey from the main study allowed us to estimate both patient and provider-side costs of delivering TB diagnostic and treatment services.

For the BIA analysis, we extracted data on TB diagnostic service utilization and treatment initiation within 7 days, asset ownership, and socio-economic status across the trial arm and each country from the TB-CAPT core trial survey. From the patient cost survey, using structured survey tool we gathered data on direct medical expenses (such as consultation fees, chest X-rays, laboratory and medication fee, and other supplies), direct non-medical costs (including travel and additional food), and indirect costs related to time spent and lost working days while seeking care. Indirect costs were adjusted using 2023 minimum wage estimates for each country [20]. Similarly, for the health system cost survey, we analyzed provider cost information related to TB testing and procedures. This included expenditures on equipment, personnel, consumables, training, communication, monitoring and evaluation, and other TB diagnostic-related services. We conducted an ingredients-based cost analysis separately for Mozambique and Tanzania to estimate facility-based diagnostic costs per test in each trial arm [21]. Both patient and health system costs were converted to United state Dollar (USD) using the average exchange rate during the study period (1 USD = 2,505 Tanzanian Shillings, 63.25 Mozambican Metical) from August 2022 to June 2023 [22].

### Analysis

#### Wealth index construction

We developed a wealth index measure to assess the benefits across wealth distributions for each participant groups. The wealth index was constructed using variables related to asset ownership, which is captured through the Equity Tool. The Equity Tool contains a country specific questionnaire (e.g. Presence of a refrigerator, a mobile phone, a television, electricity, type of toilet, water supply, cooking fuel, and materials of the wall and floor etc) mainly built from the full wealth index parameters from the Demographic and Health surveys [23]. We recorded the variables from the Equity Tool to prepare for multiple correspondence analysis (MCA) [24]. MCA is conceptually similar to principal component analysis used for exploring and visualizing the relationships among multiple categorical variables. The responses were recoded as 0 (absence) or 1 (presence) for binary variables and transformed categorical variables with multiple options into dummy variables, each representing a specific category coded as either 0 or 1. We then conducted MCA separately for each country using STATA software. The MCA allowed us to reduce the dimensionality of the data and capture underlying patterns in asset ownership. The analysis produced a set of dimensions, with the first dimension explaining the greatest variation in socio-economic status. Each variable received a factor score based on its contribution to this dimension. We used these scores to generate a composite wealth index for each participant, reflecting their relative socio-economic position. Participants were then ranked according to their wealth index scores and grouped into quintiles. Finally, both country-specific and combined wealth index data for both countries were used for the benefit-incidence analysis (BIA).

#### Benefit Incidence Analysis

The BIA combined information on the health benefits (i.e., use of TB diagnostic and early treatment initiation) with information on the cost of providing the services to assess the incidence of the benefit from public spending across the wealth index. The aim was to understand how public spending on TB diagnostic tests is distributed across different wealth index, providing insights into equity of resource allocation.

In the first step of BIA analysis, the health benefit or outcome measure was defined as the initiation of TB treatment within 7 days of diagnosis, consistent with the TB - CAPT core trial main outcome. We created a binary variable indicating whether each participant-initiated TB treatment within 7 days of diagnosis. For each wealth quintile, we calculated the treatment initiation rate by determining the proportion of participants who initiated treatment within this timeframe among those accessed diagnostic tests. In the second step of BIA, we calculated the unit cost of providing TB diagnosis and treatment initiation per participant in both the intervention and control arms. Specifically, we estimated the health system cost of the intervention and control arm from the health facility survey data for each country [25]. In addition, as costs where not disaggregated, the health system unit cost where assumed to remain constant across country and trial arm. The health system perspective includes all costs borne by healthcare providers (public or private) to deliver services. In addition to the health system perspective, we employed a comprehensive BIA approach including the patient cost along with the health system cost in the analysis to assess the total financial benefits of decentralization of the point-of-care diagnostic tool across wealth quintiles. Such addition offers a more detailed assessment of the financial benefit distribution, particularly those in lower wealth quintiles. Lastly, we estimated the public subsidy (total) benefit per quintile by multiplying the treatment initiation rate by unit cost of each TB diagnostic strategy [26]. The BIA analysis was performed with and without the inclusion of TB patient costs. This approach provided insights into how each TB diagnostic strategy benefits (under both provider-only cost and comprehensive scenarios) are distributed across different wealth quintiles, reflecting the financial investment provided for individuals undergoing TB diagnosis and treatment.

The TB specific benefit received by individual (*i) is estimated as S*_*ki*_ = *q*_*ki*_ *c*_*ki*_

Where,

*q*_*ki*_ is treatment initiation rate of k for individual I (i.e. TB diagnosis and treatment initiation within 7 days),

*c*_*ki*_ is the unit cost of k for individual I (i.e., health system cost with or without TB patient cost).

NB: For estimating subsidy (total) benefits, we defined beneficiaries as individuals who were diagnosed and initiated TB treatment within 7 days. Individuals who were tested but not diagnosed with TB, or who did not initiate treatment within the defined timeframe, were assigned a treatment initiation rate of zero (*q*_*ki*_ = 0), and thus were not counted as benefit recipients in this analysis.

Furthermore, to graphically assess the economic inequality in the benefit of TB treatment initiation within 7 days, we used a concentration curve and index. A concentration curve plots the cumulative proportion of the population benefiting from the TB subsidy (total benefit) against the cumulative proportion ranked by wealth index. If the curve overlaps with the line of equality, it indicates equitable distribution. A curve above (below) the line of equality suggests a pro-poor (rich) concentration of subsidy or total benefit [27]. The concentration index summarizes the magnitude of socioeconomic-related inequality in TB treatment initiation within 7 days and positive (negative) values indicate a concentration of the indicator among the advantaged (disadvantaged) groups, while zero value indicates the absence of inequality [27]. Additionally, a Kolmogorov-Smirnov test is employed to assess the statistical significance between the concentration curves, with a p-value of less than 0.05 indicating statistical significance.

### Ethical Approval and consent to participate

An ethical approval was obtained from regulatory and ethical bodies in Tanzania (National IRB approval #NIMR/HQ/R.8c/Vol.I/2323 and TMDA approval #BD.59/62/46/05) and Mozambique (National IRB approval #217/CNBS/22).

## Results

### Distribution of TB diagnosis and treatment initiation within seven days

The proportion of TB treatment initiation within 7 days among those who received diagnostic tests, in both Mozambique and Tanzania, showed a declining trend from the poorest (Q1) to the least poor quintile (Q5). In Mozambique, treatment initiation rates were highest among individuals in the poorest quintile (Q1) is 7.3% and decreased steadily across quintiles, reaching 4.5% in the poor quintile (Q4), before rising slightly to 6.2% in the least poor quintile (Q5). A similar trend was observed in Tanzania, where the poorest quintile (Q1) had the highest initiation rate at 8.4%, followed by a consistent decline across quintiles, dropping to 3.8% in the least poor quintile (Q5).

In Mozambique, the distribution of treatment initiation rate was 25% in the poorest quintile, compared to 18% in the least poor quintile. Similarly, in Tanzania, the treatment initiation rate was 27% in the poorest quintile, compared to 12% in the least poor quintile. The table below presents wealth and geographic related inequalities in TB treatment initiation within 7 days of diagnosis, showing that individuals in poorer quintiles are more likely to receive early treatment (Table 1). Furthermore, the TB diagnosis rates were higher among poorer quintiles in both Mozambique and Tanzania, reflecting a greater disease burden. However, treatment initiation within 7 days was consistently lower among wealthier groups. In Tanzania, 78% of diagnosed individuals in Q1 started treatment promptly, compared to 48% in Q5, with a similar pattern in Mozambique. This suggests that timely treatment is influenced not just by TB prevalence but also by socioeconomic factors that may delay care among higher-income quintile.

**Table 1.** Proportion of TB diagnosis and treatment initiation within 7 days by quintile, country, TB-CAPT CORE trial, 2022-2023.

| Country | Category | Quintile |  |  |  |  |
| --- | --- | --- | --- | --- | --- | --- |
|  |  | Q1<br>(Poorest) | Q2 (less<br>poor) | Q3<br>(Middle) | Q4<br>(Poor) | Q5 (least<br>poor) |
| Mozambi<br>que | Diagnosed with TB | 34 | 34 | 47 | 42 | 42 |
|  | Initiated treatment <7 days | 28 | 23 | 22 | 19 | 20 |
|  | % (treatment/ Diagnosis) | 82% | 68% | 47% | 45% | 47% |
|  | % (treatment/ Total N) | 7.3% | 6.1% | 5.5% | 4.5% | 6.2% |
|  | Total N | 385 | 377 | 397 | 418 | 323 |
| **Tanzania** | Diagnosed with TB | 45 | 45 | 42 | 34 | 31 |
|  | Initiated treatment <7 days | 35 | 34 | 26 | 20 | 15 |
|  | % (treatment/ Diagnosis) | 78% | 76% | 62% | 59% | 48% |
|  | % (treatment/ Total N) | 8.4% | 8.2% | 6.2% | 4.5% | 3.8% |
|  | Total N | 418 | 417 | 418 | 442 | 392 |
| **Overall** | Diagnosed with TB | 79 | 79 | 89 | 76 | 73 |
|  | Initiated treatment <7 days | 63 | 57 | 48 | 39 | 35 |
|  | % (treatment/ Diagnosis) | 80% | 72% | 54% | 51% | 48% |
|  | % (treatment/ Total N) | 7.9% | 7.2% | 5.9% | 4.5% | 4.9% |
|  | Total N | 803 | 794 | 815 | 860 | 715 |

The figure below shows treatment initiation by trial arm across quintiles (pooled across countries), emphasizing the intervention effect (Figure 1). The TB treatment initiation within 7 days showed differences between the intervention (Truenat) and control (SOC) arms across quintiles. In the intervention arm over 50% of TB treatment initiation benefit concentrated in the lowest two wealth quintiles. Similarly, in the control arm over 45% of TB treatment initiation benefit concentrated in the lowest two quintiles. The Intervention group consistently showed higher TB treatment initiation rates than the control group, highlighting Truenat effectiveness in promoting early diagnosis and timely treatment initiation.

**Figure 1.**
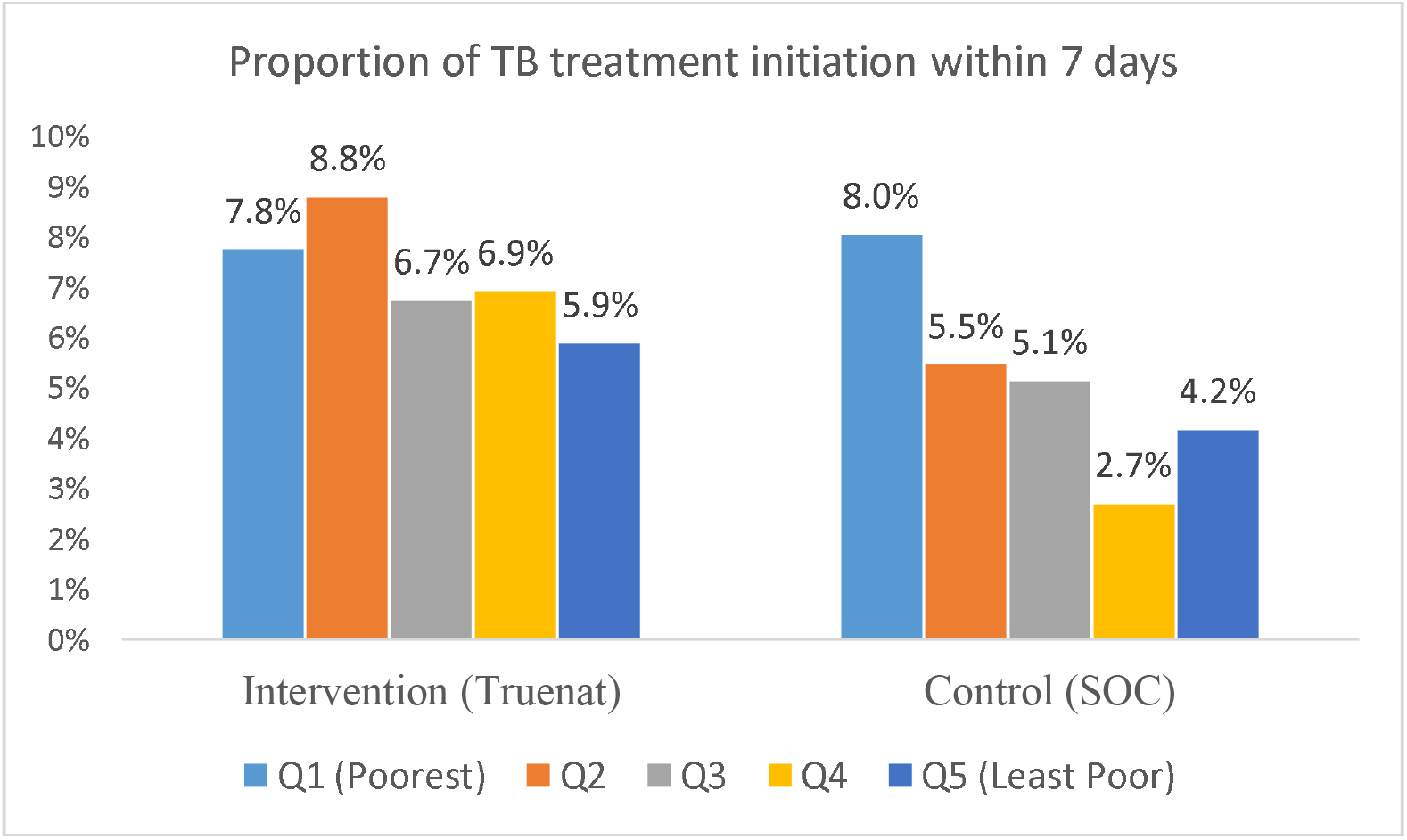
Proportion of TB diagnosis and treatment initiation within 7 days by trial arm and quintile, TB-CAPT CORE trial, 2022-2023.

The table below illustrates TB treatment initiation within 7 days by trial arm and country, showing higher rates of initiation in the intervention arm compared to the control arm across all quintiles, with more pronounced differences in Mozambique. In Mozambique, the intervention (Truenat) generally resulted in higher TB treatment initiation rates ranging from 5.2% to 8.8%, while the rate ranges from 3% to 5.2% across quintiles in the control arm (Table 3). In Tanzania, the intervention arm showed treatment initiation rates ranging from 4.7% to 11.1% across quintiles, while the control arm exhibited more variability, with rates ranging from 1.6% to 11%. Furthermore, in Mozambique, the proportion of TB treatment initiation distribution was more concentrated in the poorest (34%) compared to the least poor (15%) quintile, while in the control group, it was higher among the least poor (24%) compared to the poorest (8%). In Tanzania, the intervention arm showed slightly higher concentration among the poorest (22%) and the least poor (10%), whereas the control group demonstrated a higher concentration among the poorest (33%) compared to the least poor (14%) quintile (Table 3, Figure S1). The table below shows a comprehensive breakdown by country, trial arm, and wealth quintile, incorporating all key comparisons (Table 2).

**Table 2.** Proportion of TB diagnosis and treatment initiation within 7 days by country, trial arm, and quintile, TB-CAPT CORE trial, 2022-2023.

| Country | Category | Intervention (Truenat) |  |  |  |  | Control (SOC) |  |  |  |  |
| --- | --- | --- | --- | --- | --- | --- | --- | --- | --- | --- | --- |
|  |  | Q1 | Q2 | Q3 | Q4 | Q5 | Q1 | Q2 | Q3 | Q4 | Q5 |
| Mozambique | Yes | 25 | 16 | 12 | 10 | 11 | 3 | 7 | 10 | 9 | 9 |
|  | % | 8.8% | 6.9% | 6.4% | 5.2% | 7.3% | 3% | 4.8% | 4.9% | 4% | 5.2% |
|  | Total N | 284 | 231 | 188 | 191 | 151 | 101 | 146 | 209 | 227 | 172 |
| Tanzania | Yes | 16 | 20 | 14 | 16 | 7 | 19 | 14 | 12 | 4 | 8 |
|  | % | 6.5% | 11.1% | 7.1% | 8.6% | 4.7% | 11% | 5.9% | 5.5% | 1.6% | 3.4% |
|  | Total N | 245 | 179 | 198 | 185 | 148 | 173 | 238 | 220 | 257 | 237 |

**Table 3.** Average unit cost of TB diagnosis and treatment initiation within 7 days (USD) by country, quintile, TB-CAPT CORE trial, 2022-2023.

| Outcome | Category | Quintile |  |  |  |  |
| --- | --- | --- | --- | --- | --- | --- |
|  |  | Q1 | Q2 | Q3 | Q4 | Q5 |
| Health system cost | Mozambique | 49.8 | 47.7 | 46.0 | 45.8 | 46.0 |
|  | Tanzania | 30.2 | 32.6 | 31.7 | 36.4 | 30.4 |
| Health system + patient cost | Mozambique | 56.2 | 55.0 | 54.5 | 54.4 | 60.5 |
|  | Tanzania | 36.6 | 39.9 | 40.2 | 45.0 | 44.9 |
| Overall – Health system cost |  | 38.9 | 38.7 | 38.2 | 40.9 | 39.3 |
| Overall - Health system + patient cost |  | 45.3 | 45.9 | 46.8 | 49.6 | 53.8 |

### Distribution of the unit cost for TB treatment initiation within 7 days

The cost of TB diagnosis and treatment initiation within 7 days showed a decreasing trend in Mozambique, while in Tanzania, it fluctuates across quintiles. The average cost for TB treatment initiation without patient cost ranges from USD 45.8 to USD 49.8 in Mozambique and ranges USD 30.2 to USD 36.4 in Tanzania from across quintiles. The inclusion of patient costs increased the total cost across all quintiles (Table 3).

The average cost in the Truenat arm was consistently higher than in the standard of care arm across all quintiles. Specifically, the health system cost of TB treatment within 7 days was nearly twice as high in the control arm among the poorest quintile (USD 47 versus USD 24). When considering both health system and patient costs, the intervention arm still showed a higher cost burden for the poorest (USD 53.1 vs. USD 30.9 in the control arm). This suggests that the Truenat arm may involve higher-cost diagnostic options, potentially leading to improved access and timely treatment, albeit at a greater system-level expenditure (Figure 2).

**Figure 2.**
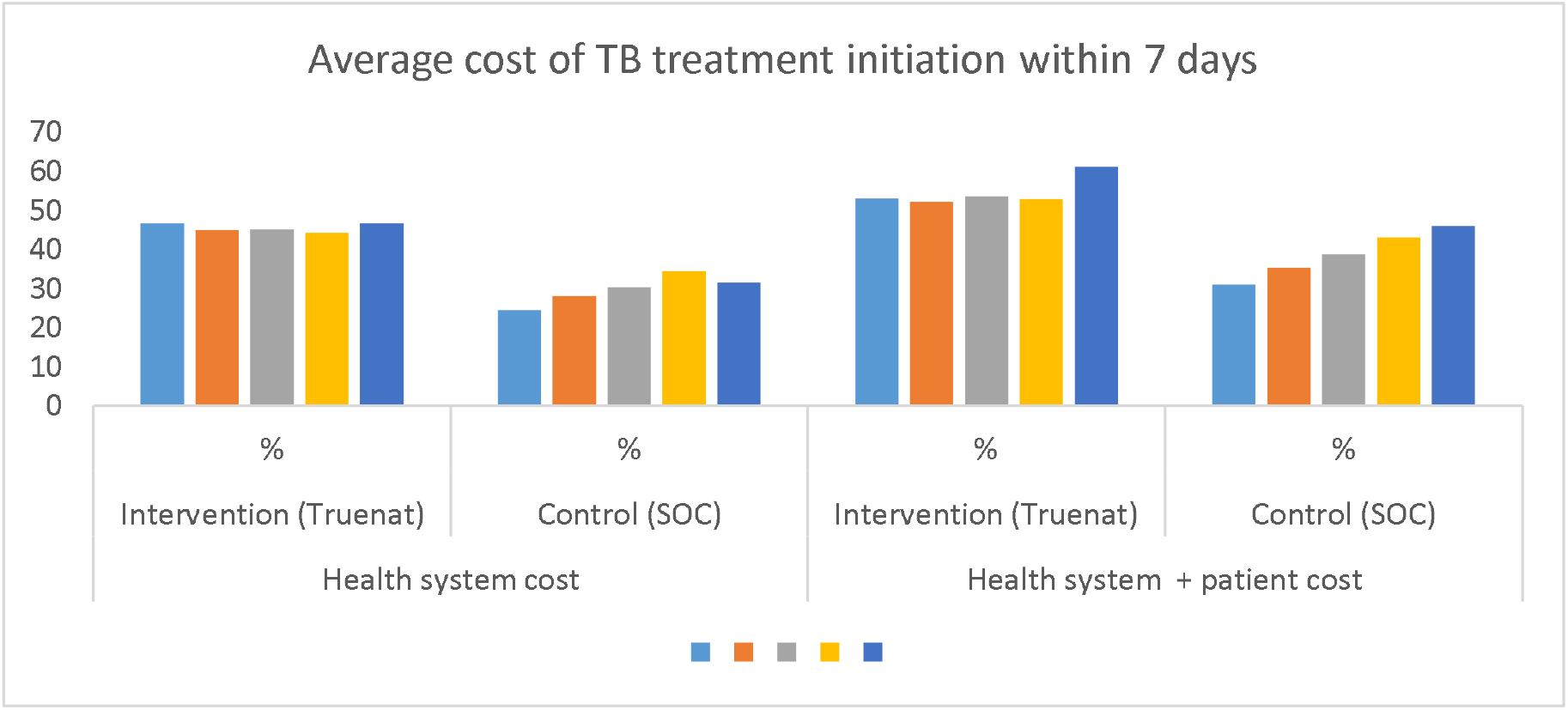
Average cost of TB diagnosis and treatment initiation within 7 days by trial arm and quintile, TB-CAPT CORE trial, 2022-2023.

The health system cost of TB diagnosis and treatment initiation was consistently higher in the intervention arm as compared to the control arm (USD 51 vs. USD 40 in Mozambique; USD 40 vs. USD 22 in Tanzania), although the cost is constant across quintiles. In Mozambique, when considering both the health system and patient costs, the values for the intervention group show an increase from USD 57.4 in Q1 to USD 65.5 in Q5, compared to the control group, which also increased trend from USD 46.4 in Q1 to USD 54.5 in Q5. Similarly, in Tanzania, when patient costs are added, the intervention group’s costs increased from Q1 (USD 46.4) to Q5 (USD 54.5), while the control group shows a lower cost ranging from USD 28.4 in Q1 to USD 36.5 in Q5. These results highlight differences in the distribution of costs between the two countries, as well as between the intervention and control groups (Table 4). Furthermore, the mean unit cost by trial arm and country with or without patient cost is depicted in annex Figure S1.

**Table 4.** Average unit cost of TB diagnosis and treatment initiation within 7 days (USD) by country, trial arm, and quintile, TB-CAPT CORE trial, 2022-2023.

| Outcome | Category | Intervention (Truenat) |  |  |  |  | Control (SOC) |  |  |  |  |
| --- | --- | --- | --- | --- | --- | --- | --- | --- | --- | --- | --- |
|  |  | Q1 | Q2 | Q3 | Q4 | Q5 | Q1 | Q2 | Q3 | Q4 | Q5 |
| Health system cost | Mozambique* | 51 | 51 | 51 | 51 | 51 | 40 | 40 | 40 | 40 | 40 |
|  | Tanzania* | 40 | 40 | 40 | 40 | 40 | 22 | 22 | 22 | 22 | 22 |
| Health system + patient cost | Mozambique | 57.4 | 58.3 | 59.5 | 59.6 | 65.5 | 46.4 | 47.3 | 48.5 | 48.6 | 54.5 |
|  | Tanzania | 46.4 | 47.3 | 48.5 | 48.6 | 54.5 | 28.4 | 29.3 | 30.5 | 30.6 | 36.5 |
| *estimates extracted from the health system cost analysis |  |  |  |  |  |  |  |  |  |  |  |

### Benefit incidence

The findings indicate that the public health system‘s investment in TB diagnostics and treatment initiation mostly benefits the poorer quintiles. The poorest quintile (Q1) receives the largest share of the benefit at 25.7%. As income rises, the share of the benefit decreases, with the least poor quintile (Q5) receiving only 16.2%. This benefit was attributed to the slightly higher treatment initiation and the higher cost of treatment subsidization by the public system. When patient costs are considered, the benefit for the poorest decreases slightly to 24.5%, while the least poor quintile‘s benefit increases to 18.2%. This suggests that patient costs reduce the benefit for the poor but still leave the public system more advantageous for them. Overall, the public investment in TB treatment was progressive but slightly less so when patient costs are included (table 5).

**Table 5.** Estimated benefit incidence of TB diagnosis and treatment initiation within 7 days by quintile, TB-CAPT CORE trial, 2022-2023.

| Outcome | N # | Quintile | TB treatment initiation rate | Average unit cost (USD) of TB treatment initiation rate | Benefit incidence share | Benefit incidence share (%) |
| --- | --- | --- | --- | --- | --- | --- |
| Benefit incidence (without patient cost) | 803 | Q1 | 7.9% | 38.9 | 3.1 | 25.7% |
|  | 794 | Q2 | 7.2% | 38.7 | 2.8 | 23.4% |
|  | 815 | Q3 | 5.9% | 38.2 | 2.3 | 19.0% |
|  | 860 | Q4 | 4.5% | 40.9 | 1.9 | 15.7% |
|  | 715 | Q5 | 4.9% | 39.3 | 1.9 | 16.2% |
|  |  |  |  |  | 11.9 |  |
| Benefit incidence (with patient cost) | 803 | Q1 | 7.9% | 45.3 | 3.6 | 24.5% |
|  | 794 | Q2 | 7.2% | 45.9 | 3.3 | 22.8% |
|  | 815 | Q3 | 5.9% | 46.8 | 2.8 | 19.0% |
|  | 860 | Q4 | 4.5% | 49.6 | 2.2 | 15.5% |
|  | 715 | Q5 | 4.9% | 53.8 | 2.6 | 18.2% |
| Total |  |  |  |  | 14.5 |  |

The concentration curve below demonstrates the overall benefit distribution of initiating TB treatment within 7 days (for both arm), situated slightly above the line of equality (Figure 3). This reflects a modestly pro-poor pattern, where the poorest groups receive a slightly larger share of the benefit compared to the least poor groups. The concentration index (CI) of –0.0816 (confidence interval: –0.0829: – 0.0804) provides further confirmation of this pro-poor benefit, indicating that early TB diagnosis and treatment initiation was more concentrated among the lower-income groups.

**Figure 3.**
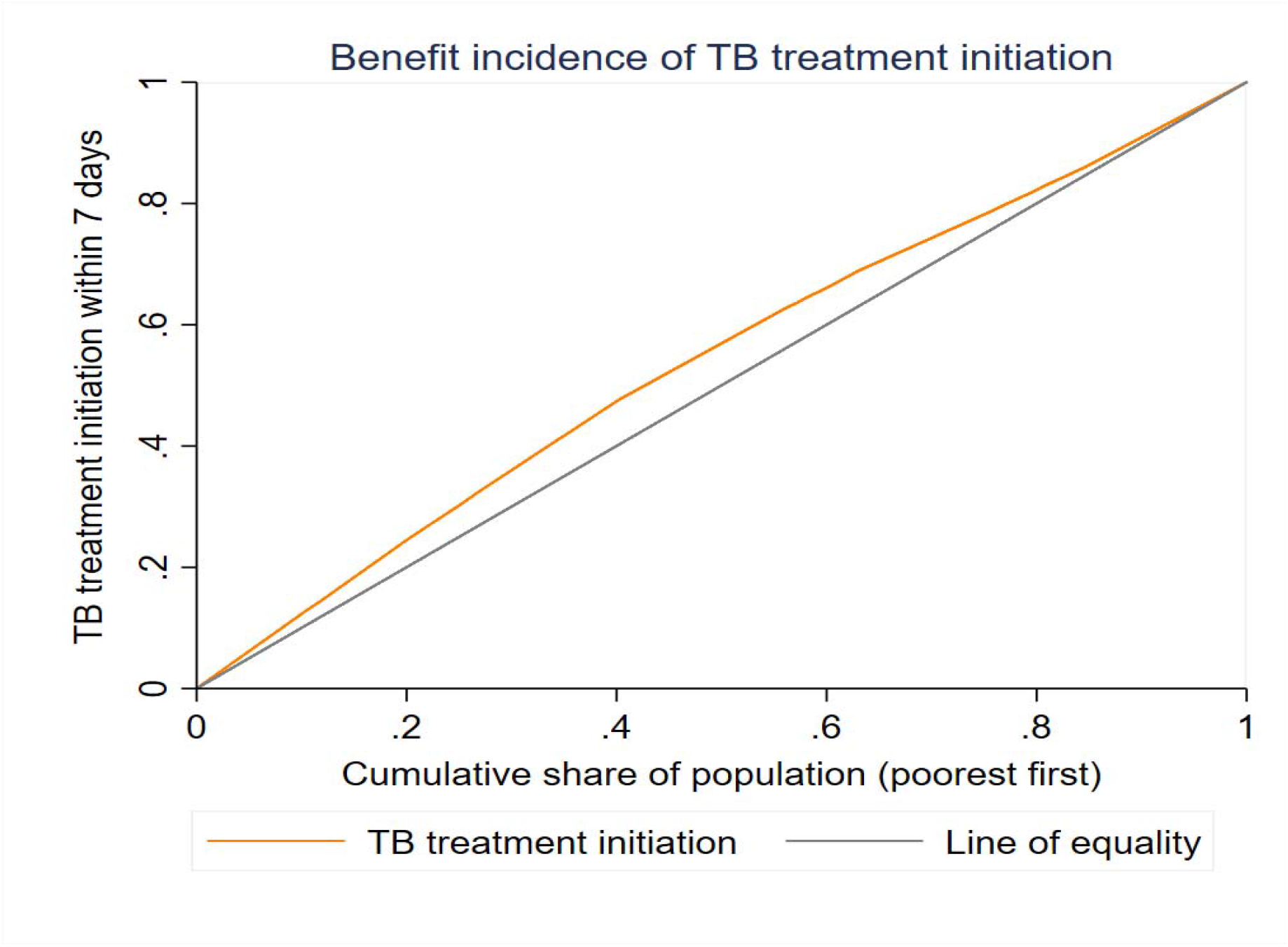
Benefit incidence of TB diagnosis and treatment initiation within 7 days (with patient cost) across quintile, TB-CAPT CORE trial, 2022-2023.

Looking at the difference between intervention and control arm, the distribution of benefit of TB treatment initiation within 7 days was slightly pro-poor for both groups. As shown in Figure 4, the concentration curves for both intervention and control arms lie above the line of equality, indicating that poorest groups receive a slightly greater benefit (Figure 4A and 4B). The CI of benefit incidence for TB treatment initiation within 7 days with patient cost for intervention and control arm is CI: – 0.0376 (confidence interval: – 0.0386: – 0.0367) and Control CI: – 0.0735 (confidence interval: – 0.0786: – 0.06844), respectively. Without patient costs, the CI for the intervention arm is – 0.0594 (confidence interval: – 0.0608: – 0.1058), and the control arm is –0.0991 (confidence interval: – 0.1029: – 0.0952). While the overall distribution of benefits remains similar with or without patient costs, there is a slight trend towards improved equity when patient costs are excluded.

**Figure 4.**
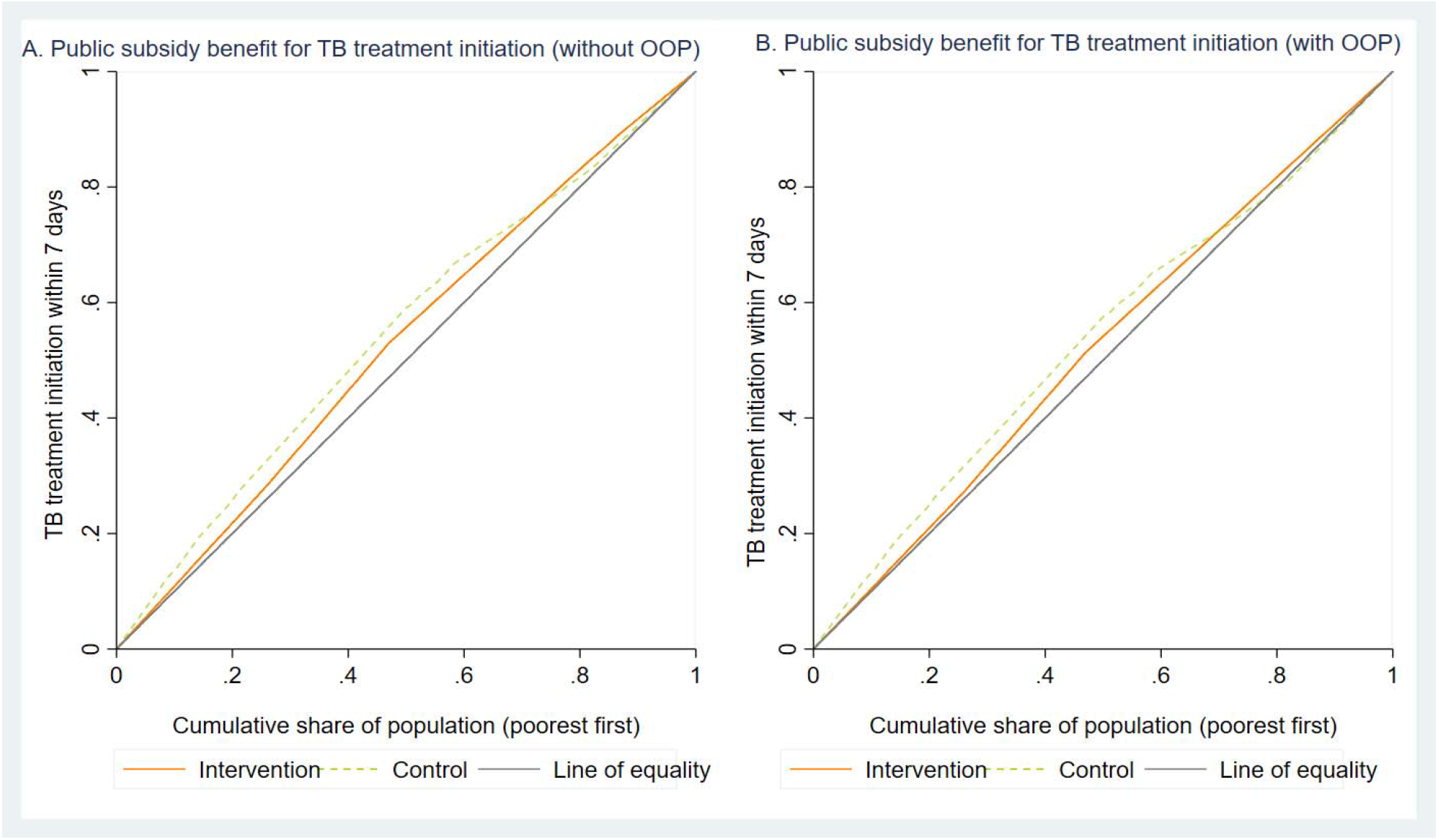
Public subsidy (total benefit) of TB diagnosis and treatment initiation within 7 days without patient costs (Figure 4A) and with patient cost (Figure 4B) across quintile and trial arm, TB-CAPT CORE trial, 2022-2023.

Furthermore, the concentration curve for the control arm lies above that of the intervention arm, the Kolmogorov-Smirnov test shows no statistically significant difference between the two groups (p > 0.05). This indicates that the distribution of treatment initiation benefits does not show a more pro-poor bias in the control group as compared to the intervention group.

The findings of the benefit incidence are largely consistent across both countries, Mozambique and Tanzania, with the benefit largely concentrated among the poorest population groups, when patient costs are excluded. In Mozambique, the intervention arm strongly benefits the poorest quintile, with this benefit declines as income levels rise. Including patient expenses slightly reduced the equity effect, but the poorest quintile still benefits more than the least poor. In contrast, the control group in Mozambique shows a more balanced distribution of benefits, with the wealthiest quintile receiving a slightly higher share, particularly when patient expenses are included. In Tanzania, the benefit incidence distribution was more equitable, with the second poorest quintile often receiving the highest share of the total benefit. The control group in Tanzania, both with and without patient cost, showed a strong pro-poor benefit, especially in the absence of patient expenses, where the poorest quintile receives the largest share of benefits (Table S2).

## Discussion

To our knowledge, this is the first time BIA has been employed to assess the distributional impact of spending made in different TB diagnostics from both the health system and patient perspectives, providing insight into how benefits are accrued by people across different SES. The use of BIA is particularly relevant for TB care, where limited resources and disparities can influence diagnostic access and treatment initiation and outcomes, transmission, and scalability of interventions. As such, our analysis can support policymakers to make evidence-based decisions about resource allocation aimed at maximizing public health benefits and its distribution in TB diagnostic services. Our findings revealed that early initiation of TB treatment after diagnosis and the distribution of public subsidies (total benefit) demonstrate a pro-poor effect, with individuals in the lowest socioeconomic quintiles benefiting the most. The consistent pro-poor benefit pattern across both diagnostic approaches suggests that both methods contribute to improving equity in access to TB diagnostics, with the intervention arm demonstrating better outcomes when it comes to prompt treatment initiation.

The benefit of TB diagnosis and treatment initiation within 7 days demonstrated a pro-poor effect, indicating that the poorest groups benefit slightly more than the least poor groups in both countries. To account for differences in TB prevalence affecting the benefit across wealth quintile, we assessed the proportion of diagnosed individuals initiating treatment within 7 days revealing that despite lower TB burden, wealthier groups had poorer linkage to care, suggesting access barriers beyond disease need. Yet, this pro-poor benefit does not differ statistically between the intervention and control arms. A study conducted in Nigeria revealed a pro-poor distribution of TB diagnostic and treatment services, with concentration indices of – 0.018 for the gross benefit and – 0.025 for the net benefit, both of which are slightly lower than the – 0.0816 found in our study. In the same study, the poorest three quintiles received 60% of the benefits from TB services [17], whereas in our study, they accounted for approximately 70% of the benefits. In general, this finding is well-documented also in other studies that interventions focused on point-of-care diagnostics at peripheral health facilities have a more pronounced pro-poor effect, as they tend to be more accessible to the poorest communities, who might otherwise face barriers in accessing diagnostic services at central or urban-focus facilities. For example, studies conducted in sub-Saharan Africa and the Asia-Pacific region have shown that rapid diagnostic tests (such as Xpert MTB/RIF) deployed in rural or peripheral settings can help overcome both geographical and financial barriers, allowing poorer populations to access timely diagnosis and treatment [9, 18, 28]. Such interventions directly address equity gaps in diagnostic access and can lead to earlier TB treatment initiation among the poor. Hence, expanding investments in point-of-care diagnostics at peripheral facilities could significantly reduce equity gaps in TB diagnoses and treatment when coupled with demand creation [17, 28, 29].

Furthermore, the inclusion of both traditional and comprehensive BIA in our study is essential, as traditional BIA reflects public subsidies alone, while comprehensive BIA accounts both health system and patient expenses to comprehensively review the distribution of benefits. Traditional BIA showed a pro-poor trend, while comprehensive BIA revealed that OOP expenses moderated this effect. This was particularly evident in the country-specific findings, for example, in Mozambique, the intervention arm provided substantial benefit in the poorest quintiles, particularly when TB patient expenses were excluded. Conversely, in the control arm, the least poor quintile benefited a slightly higher share when TB patient cost expenses were included, suggesting that financial barriers may influence the benefit distribution. In Tanzania, the benefit distribution was more equitable, with the control arm showing a stronger pro-poor bias, particularly when TB patient costs were excluded. Our dual analysis underscore the extent to which OOP expenses shape the benefit distribution across both the intervention and control arms. This aligns with broader literature, which highlights that traditional BIA focused only on public subsidies and may mask the true financial burden borne by individuals [30]. Furthermore, the impact of TB patient costs in offsetting the pro-poor impact of the benefit underscores the essential role that financial protection interventions play in making TB care equitable. Policy measures must act on reducing TB expenditure by patients with improved health financing strategies like targeted subsidies, insurance coverage, and cost-sharing exemptions [31].

The strengths of this study include the use of primary data to assess the benefit incidence of TB diagnosis and treatment initiation in high TB burden countries. However, one limitation of our study is that we defined “benefit” only for individuals who initiated TB treatment within 7 days, the analysis does not include the public spending on diagnostic services, particularly for individuals who tested negative or did not initiate treatment promptly. Moreover, the BIA was limited to the diagnostic and early treatment initiation phase; we did not assess benefit distribution across the entire TB pathway. Future studies should expand the analysis to include later stages of treatment and outcomes to assess the long-term benefit distribution. Additionally, the small sample sizes within each population subgroup and country may limit the accuracy and precision of the estimates, potentially obscuring the benefit distribution.

## Conclusion

This study demonstrates the benefit of deploying point-of-care TB diagnostics like Truenat in high-burden settings. The implementation of Truenat at peripheral facilities improved early treatment initiation and demonstrated a pro-poor distribution of benefits, particularly favoring the poorest quintiles in both countries. Importantly, the BIA reveals that the traditional approaches, which assess only public subsidies, may overstate equity gains, while the comprehensive approach with OOP inclusion appears less pro-poor. Further, our findings underscore the need for comprehensive equity assessments that incorporate both public subsidies and patient costs to better inform policy and ensure that financial barriers are addressed.

## Data Availability

The datasets used and/or analyzed during the current study are available from the corresponding author upon reasonable request.

## Acknowledgments

We acknowledge the health facilities and staffs who participated in this study, the Governments of Tanzania and Mozambique for their valued support, and the members of the TB CAPT consortium, who have been listed in a supplementary file S1, for their contribution.

