## Supplemental Appendix for "Benefit incidence analysis of decentralized Truenat MTB Plus and MTB-RIF Dx compared to hub-and-spoke Xpert MTB/RIF in Mozambique and Tanzania (TB-CAPT CORE trial)"

**Annexes**

TB treatment initiation within 7 days by country and trial arm

|  | Q1 | Q2 | Q3 | Q4 | Q5 |
| --- | --- | --- | --- | --- | --- |
| Mozambique Intervention (Truenat) | 8.8% | 6.9% | 6.4% | 5.2% | 7.3% |
| Mozambique Control (SOC) | 3% | 4.8% | 4.9% | 4% | 5.2% |
| Tanzania Intervention (Truenat) | 6.5% | 11.1% | 7.1% | 8.6% | 4.7% |
| Tanzania Control (SOC) | 11% | 5.9% | 5.5% | 1.6% | 3.4% |

Figure S1. TB treatment initiation within 7 days by country, trial arm disaggregated by quintile, by quintile, TB-CAPT CORE trial, 2022-2023.

Table S2. Estimated public subsidy benefit for TB treatment initiation within 7 days by quintile, TB-CAPT CORE trial, 2022-2023.

| Country | Outcome | Trial arm | N # | Quintile | Treatment initiation rate | Net subsidy | Subsidy benefit | Benefit incidence |
| --- | --- | --- | --- | --- | --- | --- | --- | --- |
| Mozambique | Benefit incidence (without patient cost) | Intervention | 284 | Q1 | 8.8 | 51 | 4.5 | 25.4% |
|  |  |  | 231 | Q2 | 6.9 | 51 | 3.5 | 19.9% |
|  |  |  | 188 | Q3 | 6.4 | 51 | 3.3 | 18.4% |
|  |  |  | 191 | Q4 | 5.2 | 51 | 2.7 | 15.1% |
|  |  |  | 151 | Q5 | 7.3 | 51 | 3.7 | 21.0% |
|  | Benefit incidence (without patient cost) | Control | 101 | Q1 | 3.0 | 40 | 1.2 | 13.7% |
|  |  |  | 146 | Q2 | 4.8 | 40 | 1.9 | 22.0% |
|  |  |  | 209 | Q3 | 4.8 | 40 | 1.9 | 22.0% |
|  |  |  | 227 | Q4 | 3.9 | 40 | 1.6 | 18.2% |
|  |  |  | 172 | Q5 | 5.2 | 40 | 2.1 | 24.1% |
|  | Benefit incidence (with patient cost) | Intervention | 284 | Q1 | 8.8 | 57.4 | 5.1 | 24.3% |
|  |  |  | 231 | Q2 | 6.9 | 58.3 | 4.0 | 19.5% |
|  |  |  | 188 | Q3 | 6.4 | 59.5 | 3.8 | 18.3% |
|  |  |  | 191 | Q4 | 5.2 | 59.6 | 3.1 | 15.0% |
|  |  |  | 151 | Q5 | 7.3 | 65.5 | 4.8 | 22.9% |
|  | Benefit incidence (with patient cost) | Control | 101 | Q1 | 3.0 | 46.4 | 1.4 | 12.8% |
|  |  |  | 146 | Q2 | 4.8 | 47.3 | 2.3 | 21.1% |
|  |  |  | 209 | Q3 | 4.8 | 48.5 | 2.3 | 21.6% |
|  |  |  | 227 | Q4 | 3.9 | 48.6 | 1.9 | 17.9% |
|  |  |  | 172 | Q5 | 5.2 | 54.5 | 2.9 | 26.5% |
| Tanzania | Benefit incidence (without patient cost) | Intervention | 245 | Q1 | 6.5 | 40 | 2.6 | 17.2% |
|  |  |  | 179 | Q2 | 11.2 | 40 | 4.5 | 29.5% |
|  |  |  | 198 | Q3 | 7.1 | 40 | 2.8 | 18.6% |
|  |  |  | 185 | Q4 | 8.6 | 40 | 3.5 | 22.8% |
|  |  |  | 155 | Q5 | 4.5 | 40 | 1.8 | 11.9% |
|  | Benefit incidence (without patient cost) | Control | 173 | Q1 | 11.0 | 22 | 2.4 | 40.3% |
|  |  |  | 238 | Q2 | 5.9 | 22 | 1.3 | 21.6% |
|  |  |  | 220 | Q3 | 5.5 | 22 | 1.2 | 20.0% |
|  |  |  | 257 | Q4 | 1.6 | 22 | 0.3 | 5.7% |
|  |  |  | 237 | Q5 | 3.4 | 22 | 0.7 | 12.4% |
|  | Benefit incidence (with patient cost) | Intervention | 245 | Q1 | 6.5 | 46.4 | 3.0 | 16.5% |
|  |  |  | 179 | Q2 | 11.2 | 47.3 | 5.3 | 28.7% |
|  |  |  | 198 | Q3 | 7.1 | 48.5 | 3.4 | 18.6% |
|  |  |  | 185 | Q4 | 8.6 | 48.6 | 4.2 | 22.8% |
|  |  |  | 155 | Q5 | 4.5 | 54.5 | 2.5 | 13.4% |
|  | Benefit incidence (with patient cost) | Control | 173 | Q1 | 11.0 | 28.4 | 3.1 | 38.0% |
|  |  |  | 238 | Q2 | 5.9 | 29.3 | 1.7 | 21.0% |
|  |  |  | 220 | Q3 | 5.5 | 30.5 | 1.7 | 20.3% |
|  |  |  | 257 | Q4 | 1.6 | 30.6 | 0.5 | 5.8% |
|  |  |  | 237 | Q5 | 3.4 | 36.5 | 1.2 | 15.0% |
